# Trajectory of Cognitive Function After Incident Heart Failure

**DOI:** 10.1101/2024.02.09.24302608

**Authors:** Supriya Shore, Hanyu Li, Min Zhang, Rachael Whitney, Alden L. Gross, Ankeet S. Bhatt, Brahmajee K. Nallamothu, Bruno Giordani, Emily M. Briceño, Jeremy B. Sussman, Jose Gutierrez, Kristine Yaffe, Michael Griswold, Michelle C. Johansen, Oscar L. Lopez, Rebecca F. Gottesman, Stephen Sidney, Susan R. Heckbert, Tatjana Rundek, Timothy M. Hughes, William T. Longstreth, Deborah A. Levine

## Abstract

**Background:** The size/magnitude of cognitive changes after incident heart failure (HF) are unclear. We assessed whether incident HF is associated with changes in cognitive function after accounting for pre-HF cognitive trajectories and known determinants of cognition.

**Methods:** This pooled cohort study included adults without HF, stroke, or dementia from six US population-based cohort studies from 1971-2019: Atherosclerosis Risk in Communities Study, Coronary Artery Risk Development in Young Adults Study, Cardiovascular Health Study, Framingham Offspring Study, Multi-Ethnic Study of Atherosclerosis, and Northern Manhattan Study. Linear mixed-effects models estimated changes in cognition at the time of HF (change in the intercept) and the rate of cognitive change over the years after HF (change in the slope), controlling for pre-HF cognitive trajectories and participant factors. Change in global cognition was the primary outcome. Change in executive function and memory were secondary outcomes. Cognitive outcomes were standardized to a *t-*score metric (mean [SD], 50 [10]); a 1-point difference represented a 0.1-SD difference in cognition.

**Results:** The study included 29,614 adults (mean [SD] age was 61.1 [10.5] years, 55% female, 70.3% White, 22.2% Black 7.5% Hispanic). During a median follow-up of 6.6 (Q1-Q3: 5-19.8) years, 1,407 (4.7%) adults developed incident HF. Incident HF was associated with an acute decrease in global cognition (-1.08 points; 95% CI -1.36, -0.80) and executive function (-0.65 points; 95% CI -0.96, -0.34) but not memory (-0.51 points; 95% CI -1.37, 0.35) at the time of the event. Greater acute decreases in global cognition after HF were seen in those with older age, female sex and White race. Individuals with incident HF, compared to HF-free individuals, demonstrated faster declines in global cognition (-0.15 points per year; 95% CI, -0.21, -0.09) and executive function (-0.16 points per year; 95% CI -0.23, -0.09) but not memory ( -0.11 points per year; 95% CI -0.26, 0.04) compared with pre-HF slopes.

**Conclusions:** In this pooled cohort study, incident HF was associated with an acute decrease in global cognition and executive function at the time of the event and faster declines in global cognition and executive function over the following years.

**Clinical Perspective:** *What is new?:*

- Incident heart failure (HF) is associated with an acute decrease in global cognition and executive function at the time of the event and also faster declines in global cognition and executive function during the years after the event, controlling for pre-HF cognitive trajectories.

*What are the clinical implications?:*

- Preventing HF might be an effective strategy for maintaining brain health.
- Cognition should be assessed after HF diagnosis.
- HF management should be tailored to cognitive abilities.

## INTRODUCTION

Understanding and reducing the vascular contributions to cognitive decline and dementia is critical because dementia is common, costly, and morbid. Heart failure (HF) affects 6.2 million people worldwide and imposes a high burden of disabling symptoms on afflicted individuals, associated caregivers, healthcare systems, and society.^1, 2^ Management of HF is complex, relying heavily on a patient’s cognitive ability due to the need for significant care coordination, lifestyle changes, medication compliance, and to process multi-faceted information about available therapies.^3–5^ While cognitive dysfunction has been linked to several cardiovascular disorders like hypertension, coronary artery disease, and stroke,^6–9^ how cognition changes after incident HF remains poorly understood.

Prior studies note a high prevalence of cognitive impairment in HF patients.^10–12^ However, the size of acute changes in cognition at the time of incident HF is unclear. It is also unclear whether HF survivors have a faster rate of cognitive decline (slope change) over the years following HF onset compared with their pre-HF rate of cognitive decline after accounting for the acute cognitive decrease at the time of the event (intercept change). Studies describing changes in cognition after HF have been unable to measure actual changes in cognition associated with HF because they lacked repeated measures of cognition before and after HF onset.^13–15^.Additionally, studies have not measured both the acute decrease in cognition at the time of HF onset and the change in the rate of cognitive decilne over the years after HF simultaneously. As the number of people with HF is expected to increase,^16^ identifying how cognition changes during the disease course has significant implications for clinical practice and healthcare policy.^17^

We conducted a pooled study using six population-based cohorts with repeated cognitive testing, data on incident HF, and longitudinal follow-up to determine the changes in cognition after incident HF. We hypothesized that incident HF is associated with an acute decrease in cognition and a faster rate of cognitive decline, after accounting for pre-HF cognitive trajectories. If an association was found, we also aimed to assess if the association between HF and cognitive change varied by age, sex and race.

## METHODS

### Data Source and Study Design

This cohort study included individual participant data from 6 well-characterized, US prospective cohorts and study years between 1971 and 2019: Atherosclerosis Risk in Communities Study (ARIC), Coronary Artery Risk Development in Young Adults Study (CARDIA), Cardiovascular Health Study (CHS), Framingham Offspring Study (FOS), Multi-Ethnic Study of Atherosclerosis (MESA) and Northern Manhattan Study (NOMAS). These cohorts were chosen because they have physician-adjudicated HF cases, repeated measures of cognition and blood pressure (BP), and geographic, age, racial or ethnic diversity.^18–22^

### Ethics

The institutional review board at the University of Michigan approved this study. Participating institutions approved the cohort studies. All participants gave written informed consent. Data will be shared according to the procedures of the cohort studies. All analyses were pre-registered and approved by each cohort’s leadership teams prior to initiation.

### Study Population

We included all individuals with at least one cognitive assessment during cohort follow-up and without a preceding history of HF, stroke, or dementia (Figure 1). We excluded individuals with HF, stroke, or dementia at or before the first cognitive assessment and those lacking complete data on covariates of interest. Follow-up was censored after an incident stroke if it occurred during follow-up, as stroke is associated with cognitive decline.^6^

**Figure 1:**
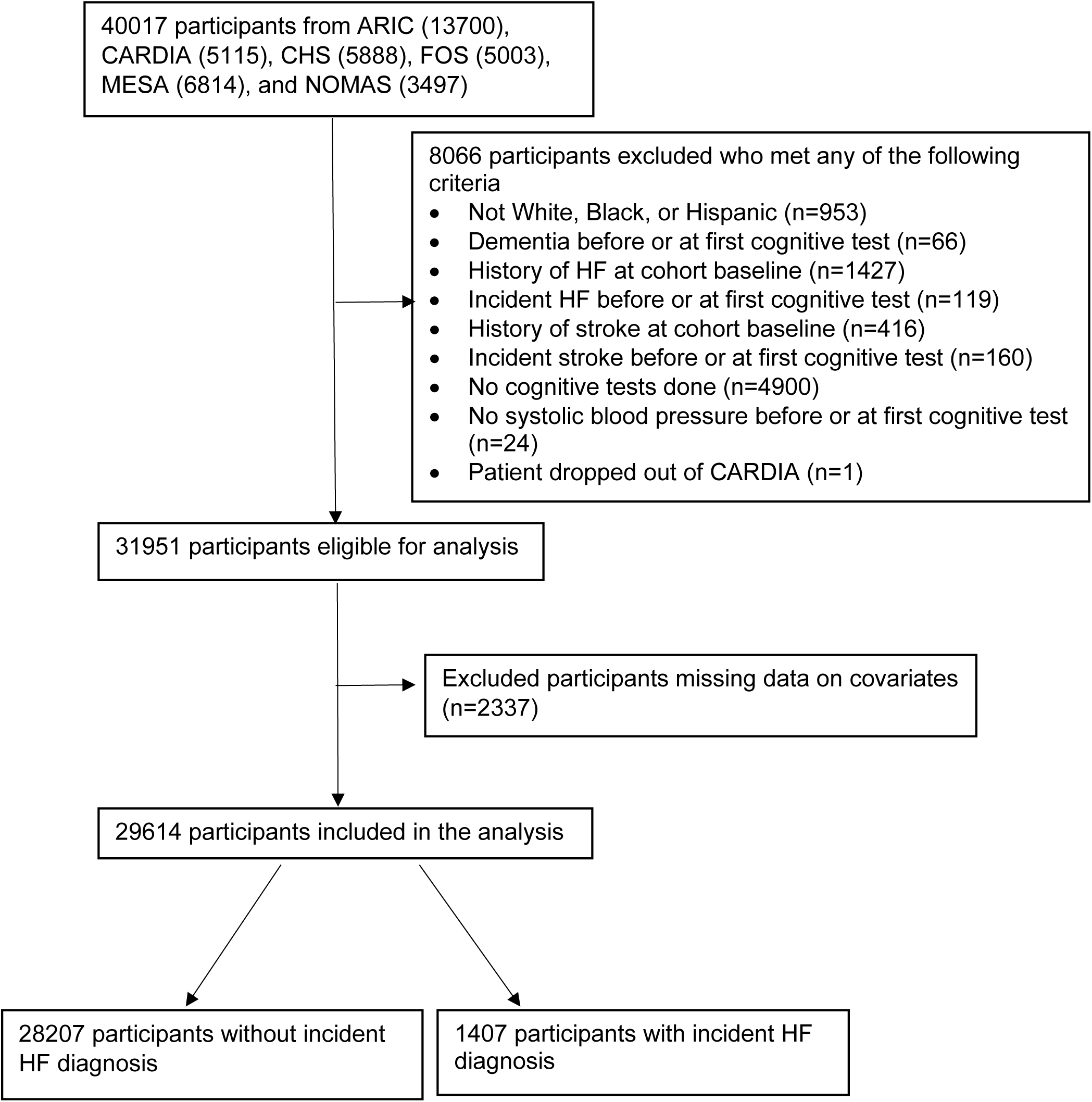
Cohort Creation Legend: ARIC: Atherosclerosis Risk in Communities Study; CARDIA: Coronary Artery Risk Development in Young Adults Study; CHS: Cardiovascular Health Study; FOS: Framingham Offspring Study; HF: Heart Failure; MESA: Multi-Ethnic Study of Atherosclerosis; NOMAS: Northern Manhattan Study

### Exposure variable

HF was captured prospectively during cohort follow-up. HF diagnoses were adjudicated by expert physicians in CARDIA, CHS, FOS, MESA, and for participants enrolled in ARIC after 2005 using standardized protocols with medical record review (see Supplementary Table 1 for details).^23–27^ For ARIC participants enrolled prior to 2005, HF diagnosis was classified as a hospitalization with an International Classification of Disease diagnosis code indicating HF in any position. For NOMAS, HF diagnosis was based on participant self-report,^28^ which also has high specificity.^29^ Measures of left ventricular ejection fraction are not available in all cohorts and were not used in our study.

### Outcome

The primary outcome of interest was global cognition (GC). Secondary outcomes were executive function (EF) and memory. Validated cognitive tests, consistent with the Vascular Cognitive Impairment Harmonization Standards,^30^ were administered in the six cohorts by trained staff either in person or over telephone.^31^

To make inferences about cognitive domains instead of individual cognitive test items and to account for different cognitive tests administered across cohorts, we cocalibrated available cognitive test items into factors representing GC (a composite of EF, memory, general mental status, language, motor and visuospatial ability), EF, and memory using all available cognitive information.^32^ These harmonized measures for cognitive function facilitate assessment of cognitive domains rather than individual tests.^7–9^ In a pre-statistical harmonization phase, neuropsychologists identified 153 test items from 34 cognitive instruments across the cohorts and determined shared and unique items across cohorts. Neuropsychologists assigned all available cognitive test items into domains. Individual tests in each domain are listed in Supplementary Table 2. Using item response theory methods, each test item was weighted based on its correlation with other items and empirically assigned a relative location along the latent cognitive trait corresponding to its estimated difficulty.^33, 34^ We computed factor scores for each domain, which were set to a *t-*score metric (mean [SD], 50[10]) at a participant’s first cognitive assessment; a 1-point difference thus represented a 0.1-SD difference in the distribution of cognition across the 6 cohorts. Higher cognitive scores indicate better cognitive function.

### Covariates

We included covariates that could confound the association between HF and cognition and were available in all cohorts before the first cognitive assessment.^7,25^ We used covariate values measured either immediately before or on the same day as the first cognitive assessment. All included covariates were harmonized by choosing common response categories for categorical variables and converting measurements to standard units for continuous variables as done previously.^7–9^

Demographics included age, race/ethnicity, sex, education, and study cohort. Race/ethnicity was self-reported and categorized as Black, White, and Hispanic (any race) by the cohorts.^32^ All other reported races were excluded as they formed a small minority in the context of this larger study (n=953; 2.4%). Clinical variables included alcohol drinks weekly, active cigarette smoking, any physical activity, body mass index, waist circumference, history of myocardial infarction (MI), history of atrial fibrillation (AF), fasting glucose, low density cholesterol, glomerular filtration rate, use of antihypertensive medications and cumulative mean systolic blood pressure and baseline cognitive slope. Expert physicians defined the presence of MI and AF at any point from cohort enrollment until the first cognitive assessment based on published guidelines, medical record review, and standardized protocols. Current antihypertensive medication use was assessed based on medication bottles and self-report. Cumulative mean systolic blood pressure was defined as a mean of all systolic blood pressures checked at in-person visits using standard protocols and equipment.^8^ All continuous variables were centered at the median.

### Statistical Analysis

We compared baseline characteristics between patients with and without incident HF during follow-up using 2-sample *t*-tests for continuous variables and χ^2^ tests for categorical variables. Each outcome measuring cognition was assessed as a continuous variable. Incident HF was treated as a time-dependent variable.

A linear mixed-effects model was used to estimate the association between HF and cognitive change, both around the time of incident HF diagnosis and in the following years, adjusted for the covariates and baseline (pre-HF) cognitive slope. It included random effects for intercept and slope to accommodate the correlation of cognitive measures within participants over time and to allow participant-specific rates of cognitive change. We included interaction terms between time and age, sex, and race/ethnicity based on prior research.^7, 8^ The first cognitive assessment is considered as time 0 for this study.

To estimate the association between incident HF and cognition, the model included a time-varying incident HF variable (changed from 0 to 1 on the date of the first incident HF diagnosis). In addition, a spline term for years after HF was added to determine if incident HF was associated with a faster rate of cognitive decline in the years following the diagnosis. For individuals with HF, this variable was 0 before their first HF diagnosis. The coefficient for this variable represents the association of incident HF and the rate of post-HF decline in cognitive function. This study considers the first cognitive assessment after incident HF the initial change with incident HF diagnosis. We found no evidence of non-linear effects of cognition on time and the effect of HF on time.

Statistical significance for all analyses was set as *P* <0.05 (2-sided). All analyses were performed using SAS version 9.4 (SAS Institute, Cary, NC) and R (version 4.2.1).

### Sensitivity and Secondary Analyses

In sensitivity analyses, to account for attrition bias, we repeated analyses with individuals who had ≥2 cognitive assessments. We also repeated analyses within cohorts to assess for heterogeneity in associations between HF and cognitive decline.

In secondary analyses, to explore whether the association of incident HF with cognitive decline differed based on age of HF onset, we added the following interaction terms of incident HF with age as fixed effects: incident HF*age at HF onset and time elapsed since incident HF diagnosis*age at HF onset. Similar analyses were performed to assess for effect modification by sex and race.

We performed additional sensitivity analyses using myocardial infarction (MI) and atrial fibrillation (AF) as time-varying covariates to assess the impact of incident MI and AF instead of treating them as fixed covariates. These additional analyses were proposed to account for any additional changes in exposure to these variables after the first cognitive assessment, as they have been previously associated with a cognitive decline and are on the causal pathway with HF and cognitive decline.^9, 35^

## Results

Our study cohort included 29,614 participants with a mean age of 61.1±10.5 years at first cognitive assessment, 54.5% were female, 70.3% were White, 22.2% were Black, and 7.5% were Hispanic. Overall, 1,407 (4.8%) had incident HF over a median follow-up duration of 6.6 (Q1-Q3: 5-19.8) years. All participants had GC assessed. However, EF and memory assessments were introduced later and performed on 26,417 (89.2%) and 19,176 (64.8%) participants respectively. Median (Q1-Q3) number of assessments were 3(2-5) for GC, 2(2-4) for EF and 2(2-3) for memory.

### Baseline Characteristics

Table 1 presents the baseline characteristics of study participants by incident HF status. Participants who developed HF were older (68 vs. 61 years) at first cognitive assessment, less likely to have Black race (18% vs. 22%) and had lower levels of education. Participants who developed HF had a higher incidence of MI (11% vs. 4%) and AF (5% vs. 2%). Compared with participants who did not develop HF, participants with HF had a lower glomerular filtration rate, mean (SD) [68 (1) vs. 75 (19) mL/min/1.73m^2^] and a higher cumulative mean systolic blood pressure [148 (21) vs. 137 (19) mmHg] despite higher rates of antihypertensive medication use (49% vs. 31%). Participants with incident HF, compared with participants who did not develop HF, had a greater median (Q1-Q3) number of assessments for GC [8 (4-11) vs. 3 (2-5)], EF [6 (4-9) vs. 2 (2-3)] and memory [4(3-4) vs. 2(2-3)]. Participants who developed HF had lower scores for GC (-1.56 points; 95% CI -0.68 to -2.44) and EF (-2.35 points; 95% CI -1.7 to -2.96) but similar scores for memory (+0.04 points; 95% CI -1.53 to +1.61) at the study visit immediately prior to incident HF compared to those who did not develop HF.

**Table 1:**
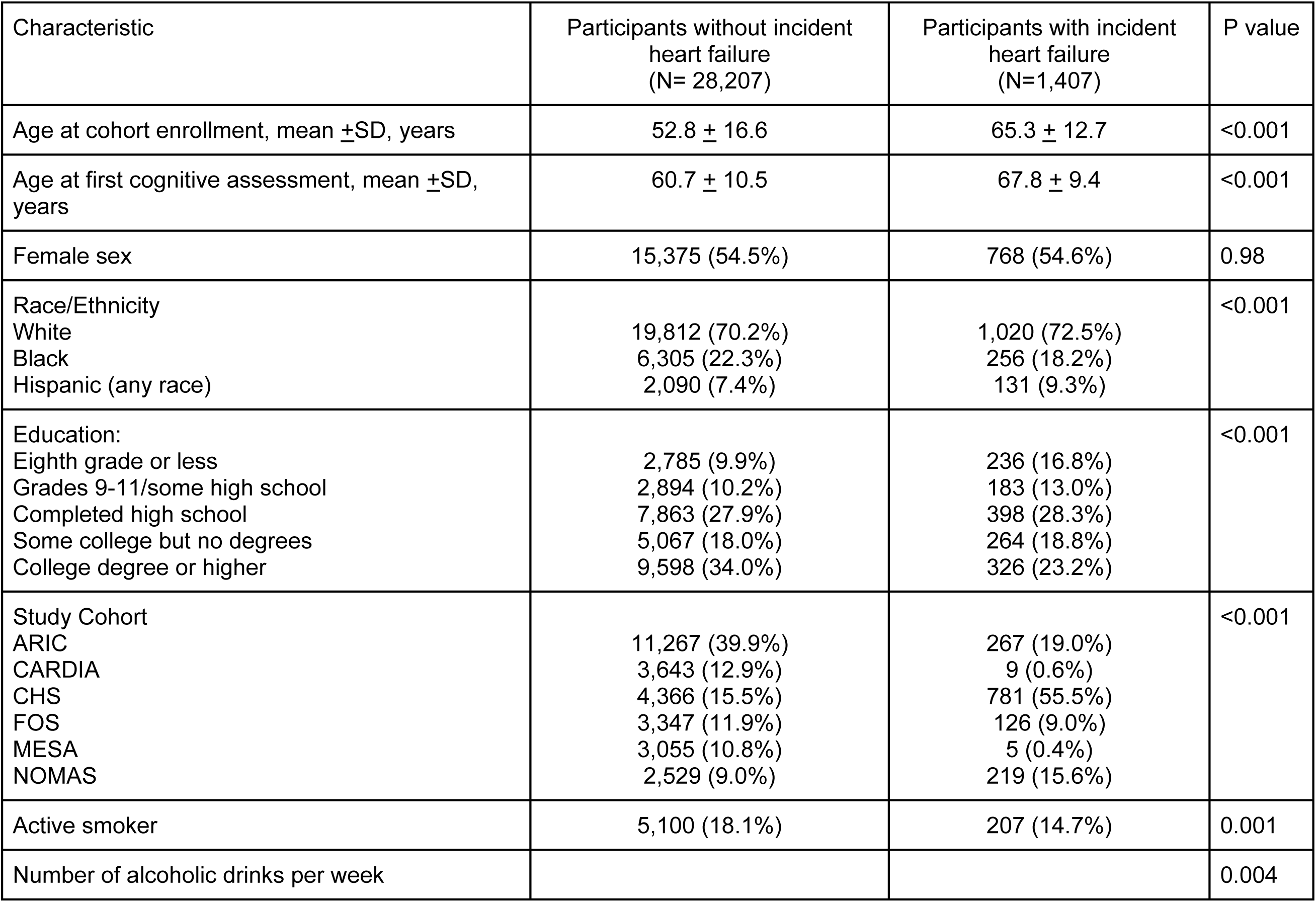
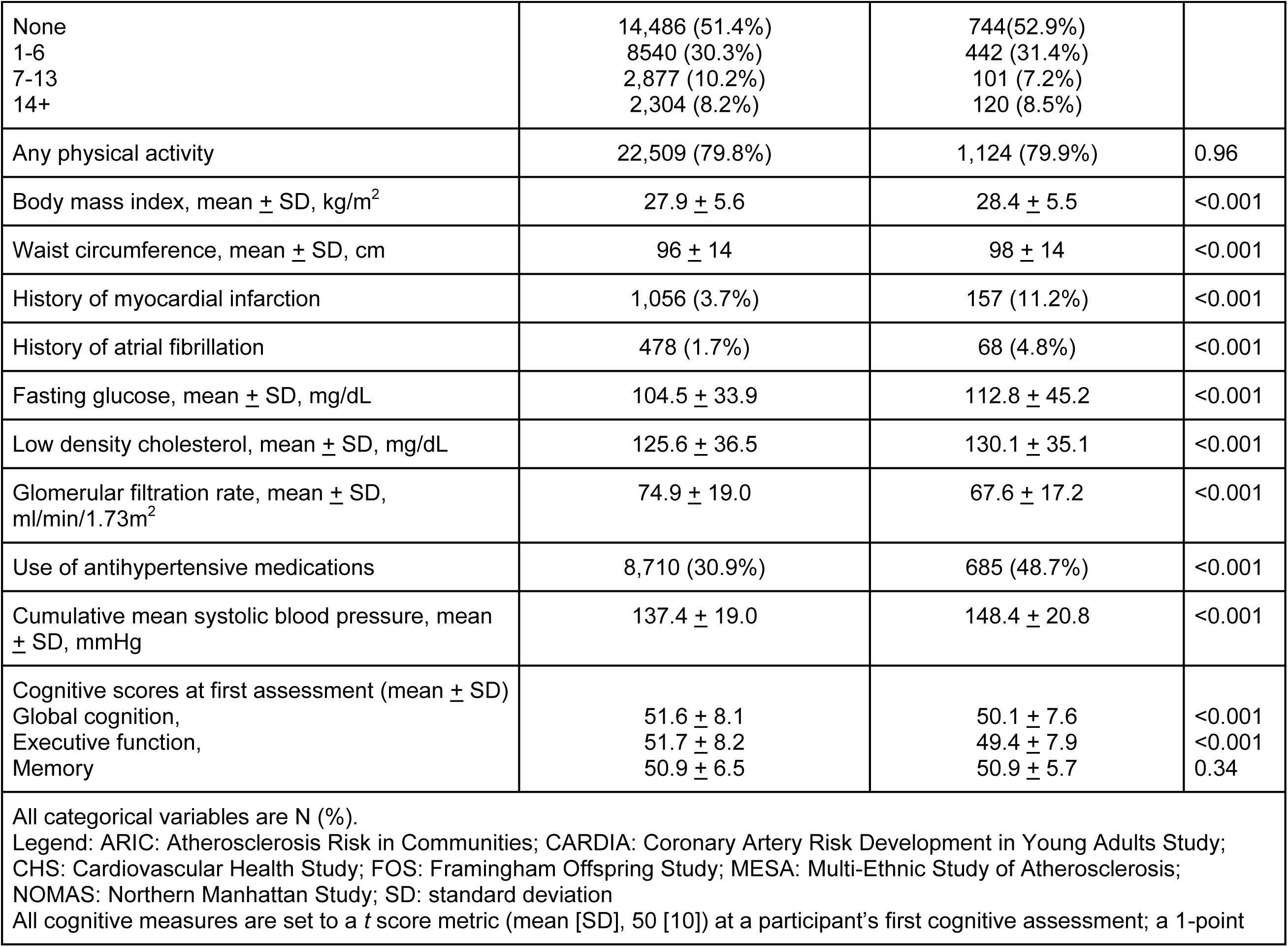
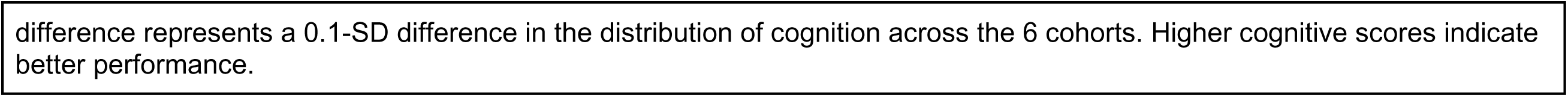
Baseline characteristics between participants who did and did not have an incident heart failure diagnosis during follow-up.

### Change in cognition with incident HF diagnosis

The median time (Q1-Q3) between the last pre-HF cognitive assessment and incident diagnosis of HF was 2.6 (1.0-6.2) years for GC, 2.2 (1.0-6.6) years for EF, and 6.1 (5.1-15.1) years for memory. After incident HF diagnosis, the median (Q1-Q3) number of assessments for GC, EF, and memory were 4 (2-8), 3 (2-6), and 2 (1-3) respectively. The median time (Q1-Q3) between incident HF diagnosis, and first post-HF cognitive assessment was 1.0 (0.9-2.2) years for GC, 1.0 (0.9-2.1) years for EF, and 5.0 (4.3-5.4) years for memory.

Incident HF was associated with an acute decrease in GC (-1.08 points; 95% CI -1.36 to - 0.80) and EF (-0.65 points; 95% CI -0.96 to -0.34) but not memory (-0.51 points; 95% CI -1.37 to 0.35) after adjusting for pre-HF slope and covariates (Table 2). Incident HF was also associated with faster decline in GC (-0.15 points per year; 95% CI -0.21 to -0.09) and EF (-0.16 points per year; 95% CI -0.23 to -0.09) but not for memory (-0.11 points per year; 95% CI -0.26 to 0.04) after controlling for pre-HF slope and the acute decrease in cognition at the time of the HF diagnosis. After selecting the final, parsimonious model, we calculated participant-specific (conditional) the predicted values for cognitive scores for a 70-year-old White male with average values of covariates at baseline (with college degree or higher education, non-smoker, no alcohol use, no history of AF, no history of MI, some physical activity, no anti-hypertensive medication use, body mass index of 27.06 kg/m^2^, waist circumference of 95.25 cm, fasting blood glucose of 97.26 mg/dL, low density lipoprotein cholesterol of 123.8 mg/dL, glomerular filtration rate 72.72 ml/min/m2, and cumulative mean systolic blood pressure of 136 mmHg.) conditional on his experiencing or not experiencing incident HF during the follow-up period (at year 9;Figure 2).

**Figure 2:**
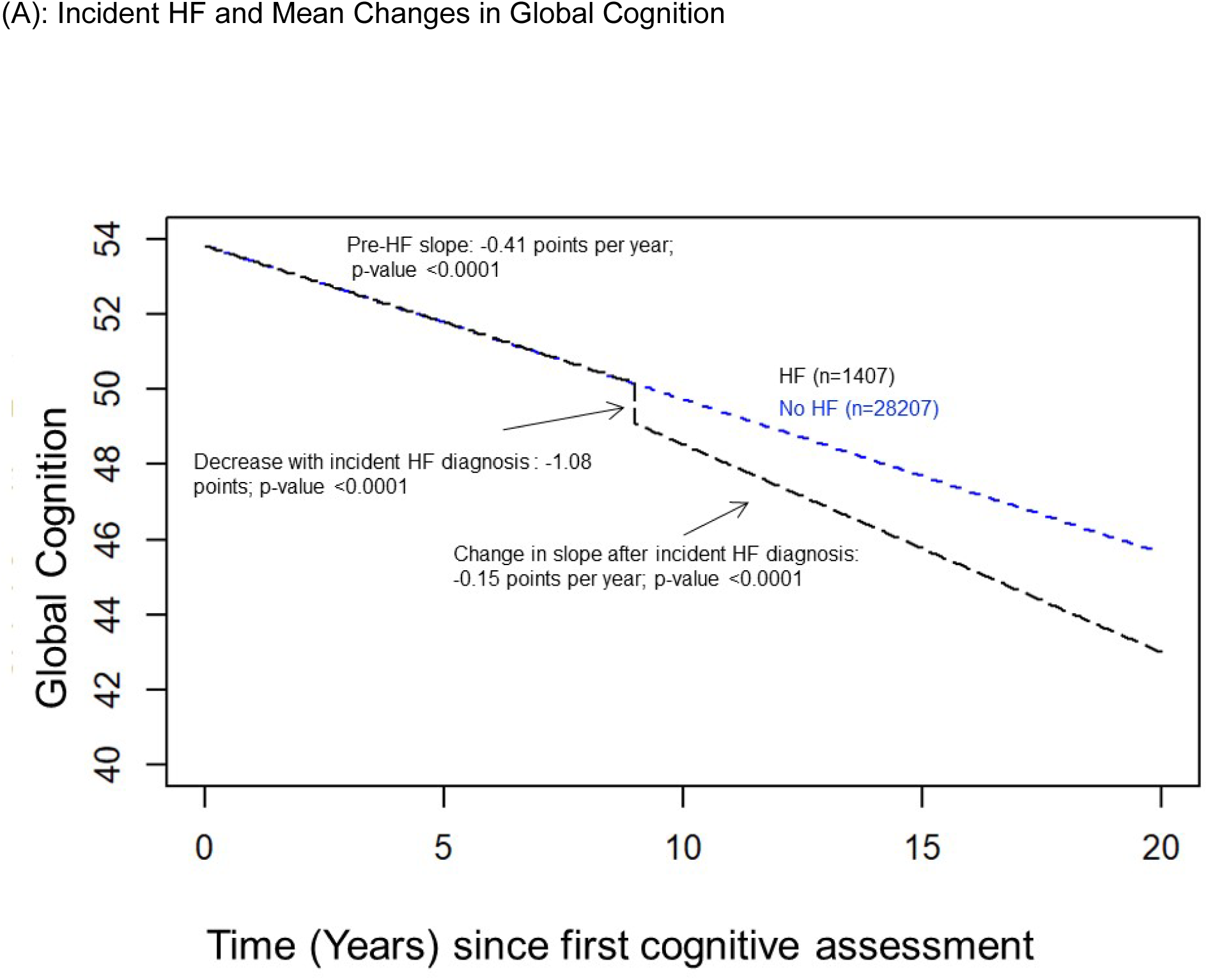
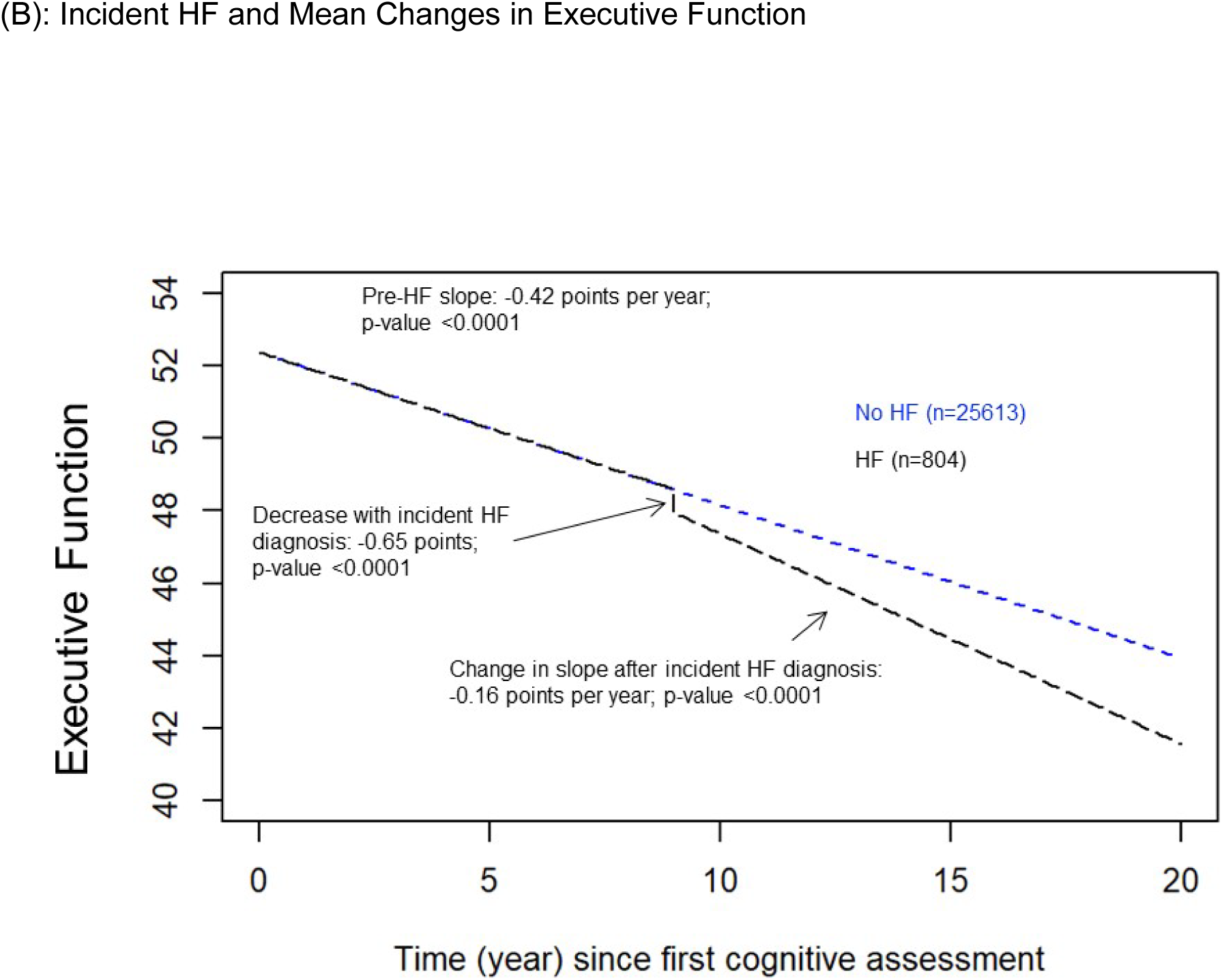
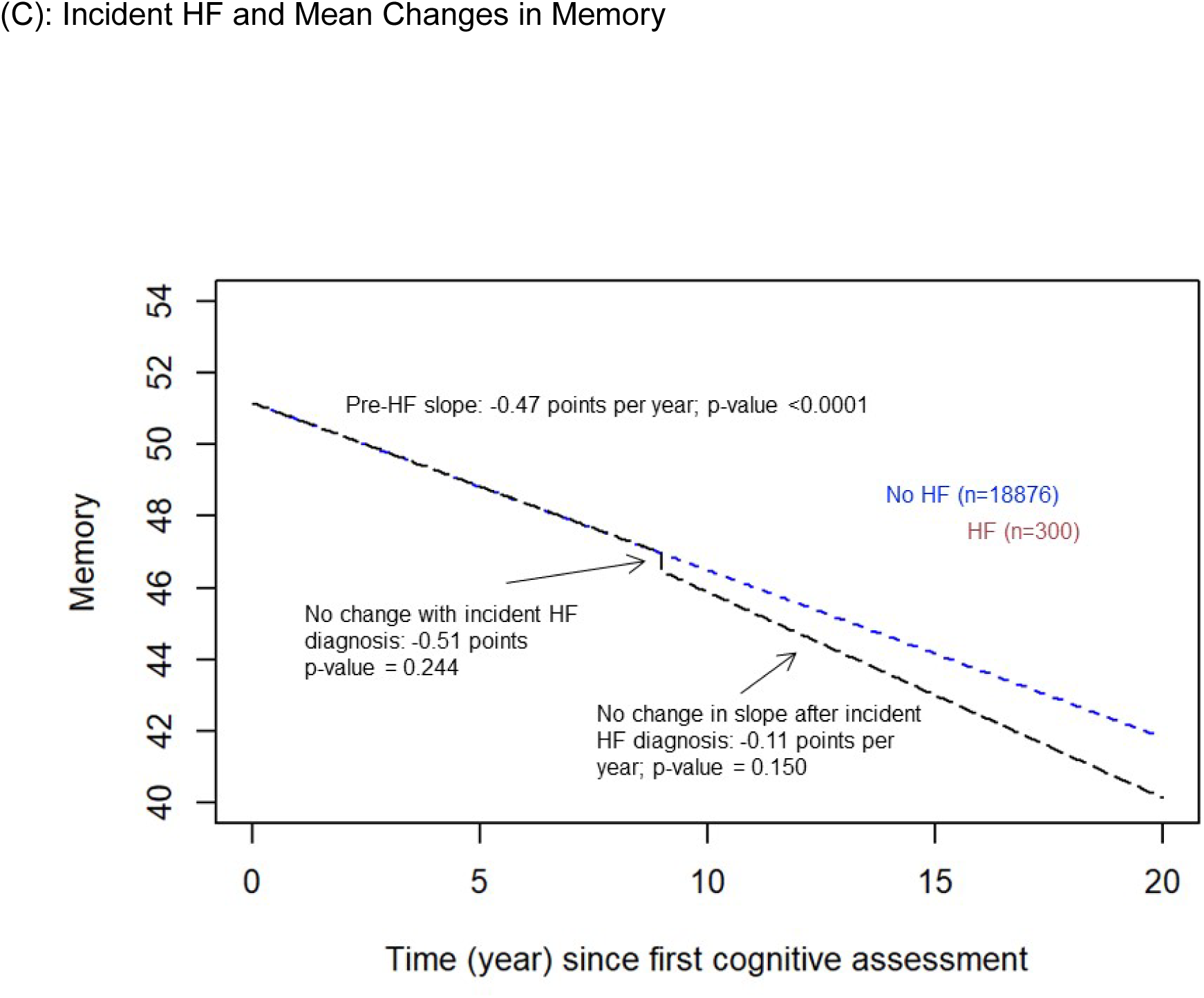
Predicted mean changes in cognitive function scores before and after heart failure diagnosis Legend: Global cognition measures global cognitive performance. All cognitive measures are set to a T-score metric (mean 50, SD 10) at a participant’s first cognitive assessment; a 1-point difference represents a 0.1 SD difference in the distribution of the specified cognitive domain across the six cohorts. Higher cognitive scores indicate better performance. Cognitive observations were censored after stroke. Graphs represent a 70-year-old White male with college degree or higher education, non-smoker, no alcohol use, no history of atrial fibrillation, no history of myocardial infarction, some physical activity, no anti-hypertensive medication use, body mass index of 27.06 kg/m^2^, waist circumference of 95.25 cm, fasting blood glucose of 97.26 mg/dL, low density lipoprotein cholesterol of 123.8 mg/dL, glomerular filtration rate 72.72 ml/min/m2, and cumulative mean systolic blood pressure of 136 mmHg. All values for continuous variables have been centered to the cohort median.

**Table 2:**
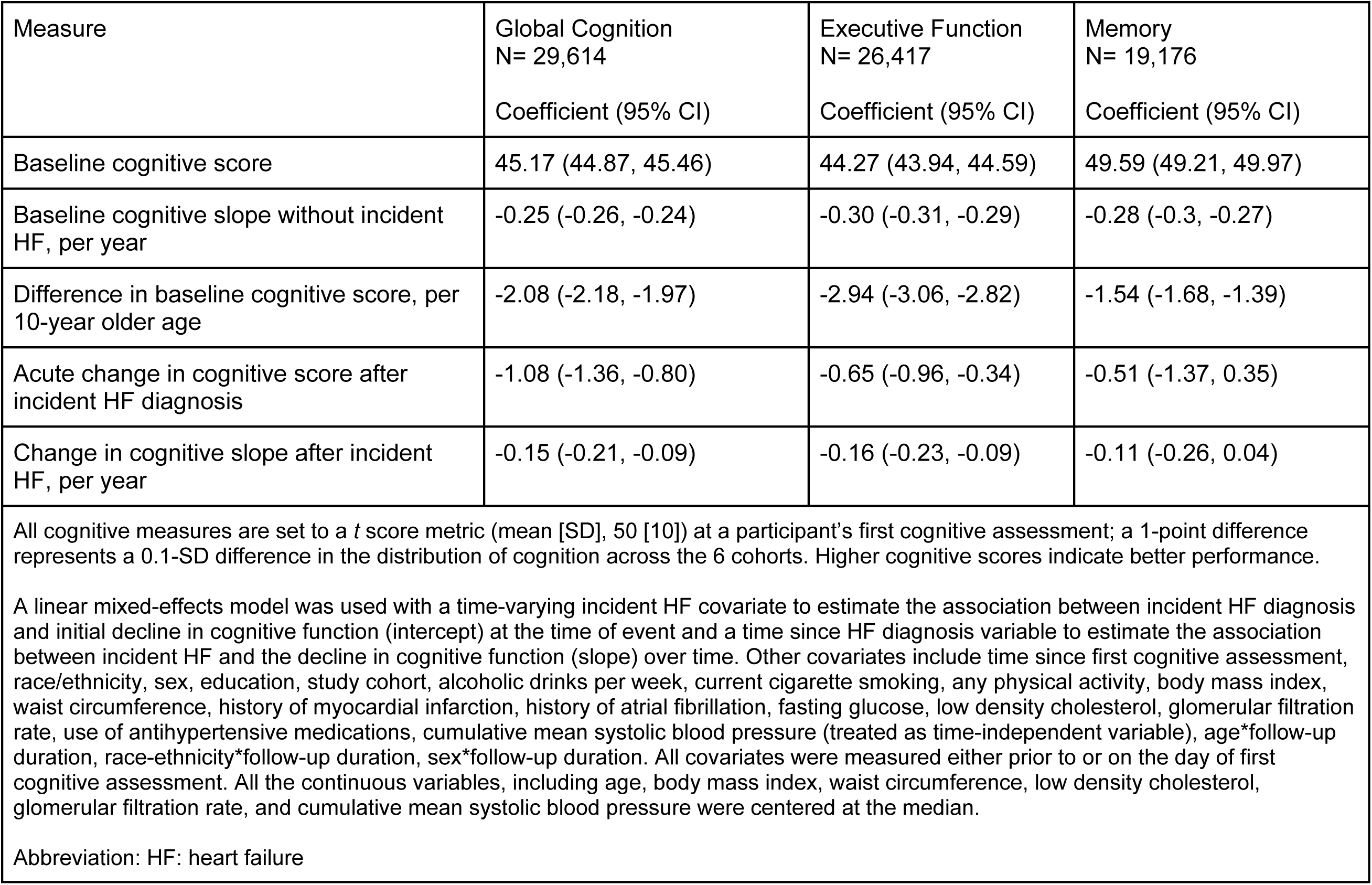
Adjusted changes in cognitive function after incident heart failure diagnosis.

| Measure | Global Cognition<br>N= 29,614 | Executive Function<br>N= 26,417 | Memory<br>N= 19,176 |
| --- | --- | --- | --- |
|  | Coefficient (95% CI) | Coefficient (95% CI) | Coefficient (95% CI) |
| Baseline cognitive score | 45.17 (44.87, 45.46) | 44.27 (43.94, 44.59) | 49.59 (49.21, 49.97) |
| Baseline cognitive slope without incident HF, per year | -0.25 (-0.26, -0.24) | -0.30 (-0.31, -0.29) | -0.28 (-0.3, -0.27) |
| Difference in baseline cognitive score, per 10-year older age | -2.08 (-2.18, -1.97) | -2.94 (-3.06, -2.82) | -1.54 (-1.68, -1.39) |
| Acute change in cognitive score after incident HF diagnosis | -1.08 (-1.36, -0.80) | -0.65 (-0.96, -0.34) | -0.51 (-1.37, 0.35) |
| Change in cognitive slope after incident HF, per year | -0.15 (-0.21, -0.09) | -0.16 (-0.23, -0.09) | -0.11 (-0.26, 0.04) |
| All cognitive measures are set to a *t* score metric (mean [SD], 50 [10]) at a participant's first cognitive assessment; a 1-point difference represents a 0.1-SD difference in the distribution of cognition across the 6 cohorts. Higher cognitive scores indicate better performance.<br>A linear mixed-effects model was used with a time-varying incident HF covariate to estimate the association between incident HF diagnosis and initial decline in cognitive function (intercept) at the time of event and a time since HF diagnosis variable to estimate the association between incident HF and the decline in cognitive function (slope) over time. Other covariates include time since first cognitive assessment, race/ethnicity, sex, education, study cohort, alcoholic drinks per week, current cigarette smoking, any physical activity, body mass index, waist circumference, history of myocardial infarction, history of atrial fibrillation, fasting glucose, low density cholesterol, glomerular filtration rate, use of antihypertensive medications, cumulative mean systolic blood pressure (treated as time-independent variable), age*follow-up duration, race-ethnicity*follow-up duration, sex*follow-up duration. All covariates were measured either prior to or on the day of first cognitive assessment. All the continuous variables, including age, body mass index, waist circumference, low density cholesterol, glomerular filtration rate, and cumulative mean systolic blood pressure were centered at the median.<br>Abbreviation: HF: heart failure |  |  |  |

### Associations between Age of Onset, Sex, and Race-Ethnicity, Incident HF and Cognitive Change

Table 3 shows the results of our interaction analyses for change in cognitive function with incident HF diagnosis by age of HF onset, sex, and race. Older age at HF onset was associated with a larger acute decrease in GC with incident HF diagnosis (-1.52 points per 10-year older age of HF onset; 95% CI -1.99 to -1.04). However, there was no difference in change in GC slope after incident HF diagnosis (0.02 points per year; 95% CI -0.05 to +0.10). While older age at HF onset did not modify change in EF acutely (-0.34 points per 10-year older age of HF onset; 95% CI -1.18, 0.50), the change in EF slope after incident HF diagnosis was slower with increasing age of HF onset (0.37 points per year per 10-year older age of HF onset; 95% CI 0.18-0.55). We found no evidence that age modified the association between incident HF and memory at either incident HF diagnosis or after.

**Table 3:**
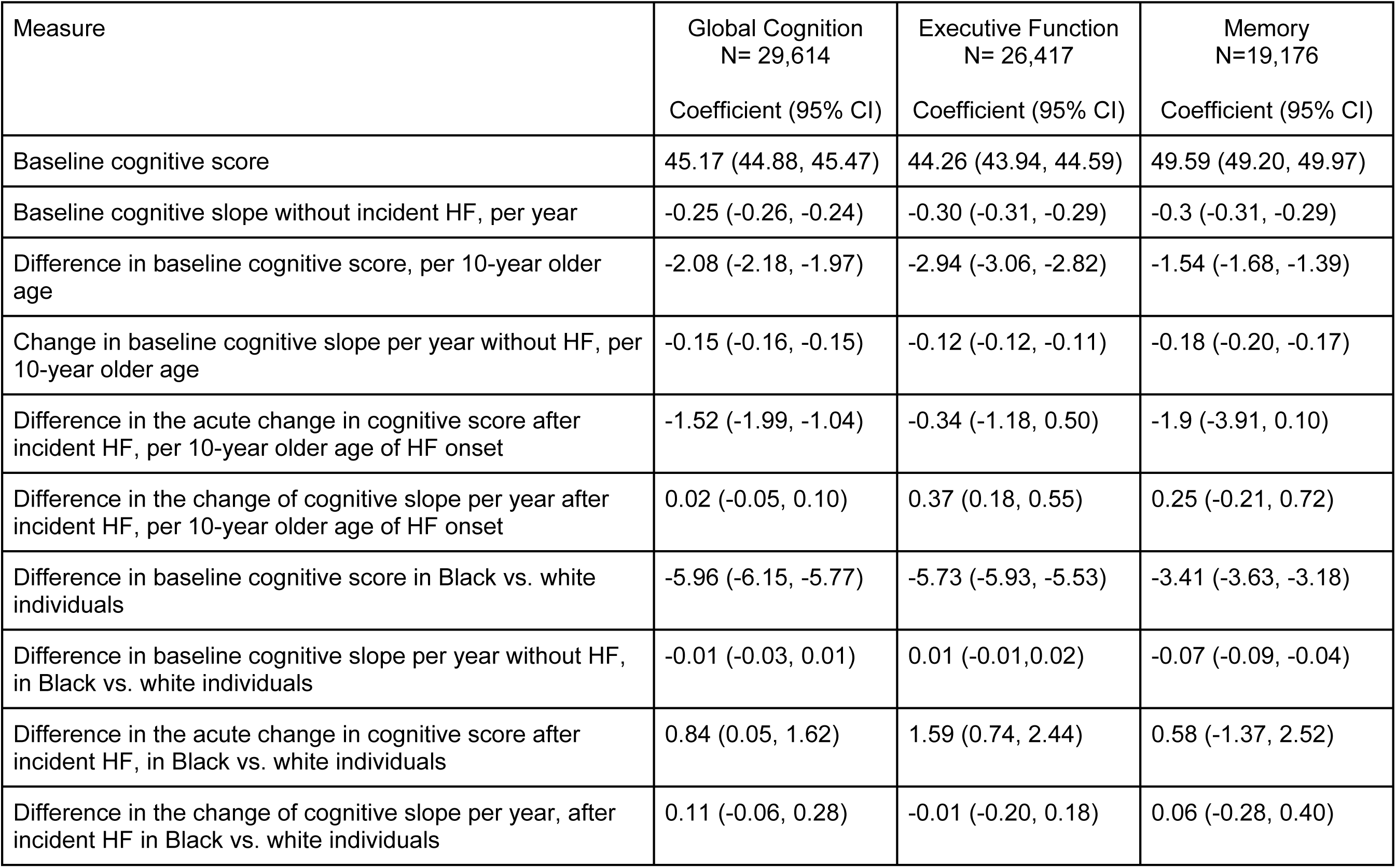
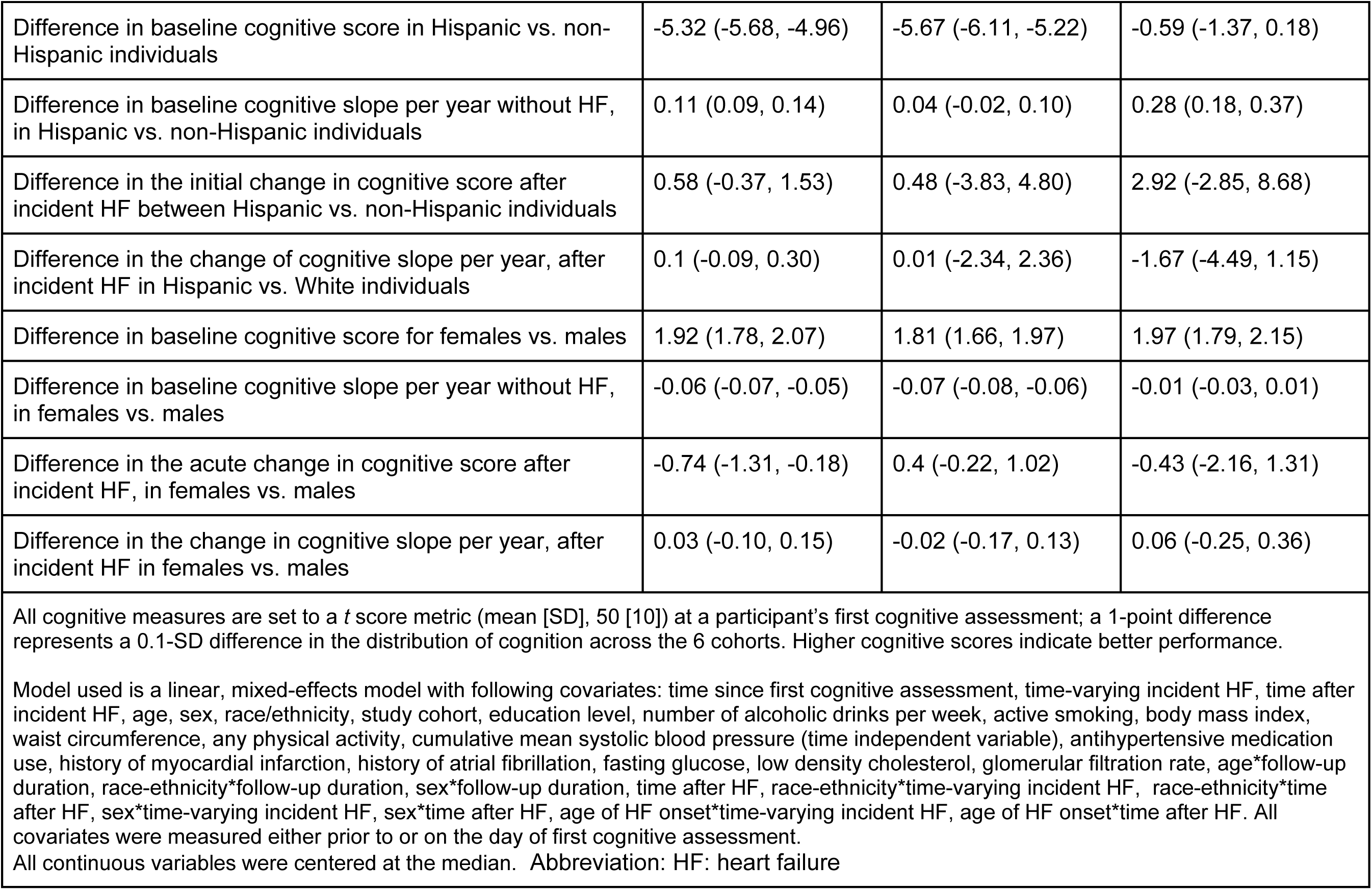
Modification of the adjusted changes in cognitive function after incident heart failure diagnosis by age of heart failure onset, race-ethnicity, and sex.

We observed racial/ethnic differences in cognitive trajectory after HF diagnosis when holding sex constant. Compared with White participants, Black participants had a smaller acute decrease in GC and EF around the time of incident HF diagnosis (0.84 points; 95% CI 0.05-1.62 & 1.59 points; 95% CI 0.74, 2.44 respectively) but not memory (0.58 points; 95% CI -1.37, 2.52). However, long-term cognitive declines after incident HF did not differ between Black and White participants for any cognitive outcome. We found no difference in the cognitive changes associated with incident HF between Hispanic (any race) and White participants.

Females experienced a larger acute decrease in GC after incident HF diagnosis (-0.74 points; 95% CI -1.31 to -0.18), but the GC slope did not differ by sex after incident HF (0.03 points per year; 95% CI -0.10 to 0.15). The EF and memory changes after incident HF did not differ by sex.

### Sensitivity Analyses

Results were similar in analyses including participants with ≥ 2 cognitive assessments (n=24,090 for GC, n=20,533 for EF, and n=14,649 for memory; Supplementary Table 3). There was little heterogeneity in the associations between HF and GC across cohorts (Supplementary Table 4). Results were similar in sensitivity analysis including history of MI and AF as time-varying covariates (Supplementary Table 5). Incident MI and AF were associated with larger acute cognitive decline in HF patients compared to those without.

## Discussion

In a pooled analysis of 6 large, US prospective cohort studies that performed serial cognitive assessments on 29,614 participants, we observed that incident HF was associated with an acute decrease in GC and EF at the time of the event as well as faster declines in GC and EC over the years after the event adjusting for pre-HF cognitive trajectories and known determinants of cognitive function. We did not find significant changes in memory associated with incident HF. Results persisted after further adjustment for MI and AF. In clinical terms, our findings suggest that individuals diagnosed with incident HF, experience global cognitive aging equivalent to 10 years within 7 years after HF diagnosis, compared to those without HF. We also observed a larger acute decrease in GC at the time of incident HF diagnosis associate with older age, female sex and self-identified White race.

Incident HF could be associated with cognitive decline through several mechanisms. We tested the best-established mechanisms, which are vascular risk factors, and comorbidities such as hypertension, MI, and AF, and they failed to explain our findings. Other hypotheses include HF causing systemic inflammation and endothelial dysfunction, which have been implicated in the causal pathway for cognitive impairment.^36–38^ In addition, HF may cause cognitive decline due to a decrease in cerebral perfusion.^39^ We likely did not see a difference in memory as we were underpowered. Additional studies need to evaluate mechanisms behind the cognitive decline associated with HF to identify modifiable targets.

Our findings suggest a larger decrease in cognition with incident HF diagnosis in older, White, and female adults. The larger acute decrease in cognition with older age likely reflects the loss of cognitive reserve and identifies subgroups at higher risk for cognitive impairment who might need closer monitoring and more support in managing HF. The larger acute decreases in GC and EF after HF seen in White participants likely reflects the higher pre-HF cognitive performance of this subpopulation^40^ with a greater room to decrease. However, sex differences in cognitive functioning are complex. Females likely have a faster cognitive decline with HF due to differences in sex hormones, greater microvascular disease, and inflammation compared to males.^7, 41, 42^

Experts have classified clinically meaningful cognitive decline as a decrease in cognitive function of ≥ 0.5 standard deviations from baseline cognitive scores.^43^ If the observed HF-related changes in cognitive function in our study are confirmed and causal, adults with HF would reach the threshold of meaningful cognitive decline 5.9 years sooner for GC and 4.7 years sooner for EF than HF-free adults. Findings from our study have several implications both for clinical management and healthcare policy. Reducing HF-associated burden is a priority.^5^ Successfully achieving this goal relies heavily on a patient’s cognitive ability with the need to follow specific diet, monitor symptoms, adhere to medications – with an average HF patient taking at least 10 different medications – and coordinate care across multiple sub-specialties.^44^ In addition, progressive cognitive decline is associated with depression,^45^ dementia,^46^ and functional decline,^47^ causing greater caregiver burden and institutionalization risk. Cognitive monitoring for older adults with HF could help identify individuals at risk for these adverse events. Such screening may also facilitate interventions such as avoiding medications contributing to cognitive dysfunction and using non-pharmacological measures such as exercise training and cognitive rehabilitation that may halt or slow cognitive decline.^48, 49^ HF treatment for older adults may need to be individualized to consider associated cognitive decline. This would involve home care programs to closely monitor these patients to prevent hospitalizations, interventions to help with medication adherence and modifying decision aids for HF treatments for patients with cognitive impairment.

Our study has several strengths. First, we used harmonized measures of cognition performed serially in participants, allowing us to control for pre-HF cognitive trajectory and describe the entire trajectory of cognitive function for HF patients. Pooling cohort studies enabled us to capture diverse populations with well-described socio-demographic characteristics and to generalize more broadly than earlier studies that were more limited in scope. Finally, HF diagnosis in our study used clinical methods and did not rely on administrative or claims measures.

Our findings should be interpreted in the context of several limitations. HF typically has an insidious onset, and diagnosis may be delayed until the progression of symptoms or a severe stage. Enrolled participants are likely to have delayed their cohort visit until they felt better following a diagnosis of HF that may have delayed capturing early acute changes in cognition. Assuming that participants’ postmortem cognitive data are missing at random might lead to immortal cohort bias. However, these factors would likely bias our findings toward underestimating cognitive decline. In addition, we used linear mixed effects models that are robust to sparse data, using all available cognitive measurements prior to HF diagnosis with additional sensitivity analyses that demonstrated consistent results. We were unable to control for possible confounders such as depression that are associated with both HF^50^ and cognitive decline^46^ or markers of HF severity such as NYHA class. Third, NOMAS relied on patient self-report to identify HF, though self-reported HF has high accuracy.^29^ Since participant reported HF is highly specific but not as sensitive as clinically validated HF diagnosis, data from NOMAS could bias our findings towards the null. Assuming that participants’ postmortem cognitive data are missing at random might lead to immortal cohort bias and underestimate cognitive declines.

In conclusion, in a pooled analysis of individual participant data from 6 large, prospective cohort studies, participants with incident HF, compared with participants without HF, had an acute decrease in GC and EF and faster declines in GC and EF accounting for pre-HF cognitive trajectories and comorbidities like stroke. A larger acute decrease in cognition after incident HF diagnosis was noted in older adults, White, and female participants. Future studies should assess the mechanisms of cognitive decline after incident HF, if specific therapies mitigate risk for cognitive decline associated with HF, and whether assessment of cognitive function should be considered as part of HF management.

## Data Availability

Data will be shared according to the procedures of the cohort studies included in this manuscript which includes ARIC, CARDIA, CHS, MESA, NOMAS and FOS

## Non-standard Abbreviations and Acronyms

AF: Atrial fibrillation
ARIC: Atherosclerosis Risk in Communities Study
CARDIA: Coronary Artery Risk Development in Young Adults Study
CHS: Cardiovascular Health Study
EF: Executive Function
FOS: Framingham Offspring Study
GC: Global Cognition
HF: Heart Failure
MI: Myocardial infarction
MESA: Multi-Ethnic Study of Atherosclerosis
NOMAS: Northern Manhattan Study
SD: Standard deviation

## Disclosures

### Funding/Support

This research project is supported by a grant R01 NS102715 from the National Institute of Neurological Disorders and Stroke (NINDS), National Institutes of Health, Department of Health and Human Service. The NINDS was not involved in the design and conduct of the study; collection, management, analysis, and interpretation of the data; preparation, review, or approval of the manuscript; and decision to submit the manuscript for publication except one representative (author RFG) of the funding agency reviewed the manuscript. The content is solely the responsibility of the authors and does not necessarily represent the official views of the National Institute of Neurological Disorders and Stroke or the National Institutes of Health. Additional funding was provided to:

Supriya Shore, MD MSCS: AHA grant ID 855105

Michelle Johanssen, MD PhD: NINDS grant K23 NS112459

Deborah A Levine, MD MPH: National Institute of Aging (NIA) grant R01 AG051827 and RF1 AG068410

Alden Gross, PhD: NIA grant K01 AG050699

Bruno Giordani, PhD: NIA Michigan Alzheimer’s Disease Research Center grant P30 AG053760

Emily Briceño, PhD: NIA P30 AG024824

### Cohort Funding/Support

The Atherosclerosis Risk in Communities Study is carried out as a collaborative study supported by National Heart, Lung, and Blood Institute contracts (75N92022D00001, 75N92022D00002, 75N92022D00003, 75N92022D00004, 75N92022D00005). The ARIC Neurocognitive Study is supported by U01HL096812, U01HL096814, U01HL096899, U01HL096902, and U01HL096917 from the NIH (NHLBI, NINDS, NIA and NIDCD). The authors thank the staff and participants of the ARIC study for their important contributions.

The Coronary Artery Risk Development in Young Adults Study (CARDIA) is conducted and supported by the National Heart, Lung, and Blood Institute (NHLBI) in collaboration with the University of Alabama at Birmingham (HHSN268201800005I & HHSN268201800007I), Northwestern University (HHSN268201800003I), University of Minnesota (HHSN268201800006I), and Kaiser Foundation Research Institute (HHSN268201800004I). This manuscript has been reviewed by CARDIA for scientific content.

The Cardiovascular Health Study (CHS) was supported by contracts HHSN268201200036C, HHSN268200800007C, HHSN268201800001C, N01HC55222, 379 N01HC85079, N01HC85080, N01HC85081, N01HC85082, N01HC85083, N01HC85086, 75N92021D00006, and grants U01HL080295 and U01HL130114 from the National Heart, Lung, and Blood Institute (NHLBI), with additional contribution from the National Institute of Neurological Disorders and Stroke (NINDS). Additional support was provided by R01AG023629, R01AG15928, and R01AG20098 from the National Institute on Aging (NIA). A full list of principal CHS investigators and institutions can be found at CHS-NHLBI.org. The content is solely the responsibility of the authors and does not necessarily represent the official views of the National Institutes of Health. The Framingham Heart Study (FHS) is a project of the National Heart Lung and Blood Institute of the National Institutes of Health and Boston University School of Medicine. This project has been funded in whole or in part with Federal funds from the National Heart, Lung, and Blood Institute, National Institutes of Health, Department of Health and Human Services, under contract No. HHSN268201500001I.

The Northern Manhattan Stroke (NOMAS) study has been funded at least in part with federal funds from the National Institutes of Health, National Institute of Neurological Disorders and Stroke by R01 NS29993.

The Multi-Ethnic Study of Atherosclerosis (MESA) was supported by contracts 75N92020D00001, HHSN268201500003I, N01-HC-95159, 75N92020D00005, N01-HC-95160, 75N92020D00002, N01-HC-95161, 75N92020D00003, N01-HC-95162, 75N92020D00006, N01-HC-95163, 75N92020D00004, N01-HC-95164, 75N92020D00007, N01-HC-95165, N01-HC-95166, N01-HC-95167, N01-HC-95168 and N01-HC-95169 from the National Heart, Lung, and Blood Institute, and by grants UL1-TR-000040, UL1-TR-001079, and UL1-TR-001420 from the National Center for Advancing Translational Sciences (NCATS). Cognitive testing at Exam 6 in MESA has been funded by grants R01HL127659 from the National Heart, Lung, and Blood Institute (NHLBI), and R01AG054069 from the National Institutes on Aging. The authors thank the other investigators, the staff, and the individuals of the MESA study for their valuable contributions. A full list of participating MESA investigators and institutions can be found at http://www.mesa-nhlbi.org.

### Disclosures

The authors have no relevant conflicts of interest.

## Notes

### Competing Interest Statement

The authors have declared no competing interest.

### Author Declarations

The institutional review board at the University of Michigan approved this study. Participating institutions approved the cohort studies.

## References

1. Roger VL, Sidney S, Fairchild AL, Howard VJ, Labarthe DR, Shay CM, Tiner AC, Whitsel LP, Rosamond WD and American Heart Association Advocacy Coordinating C. Recommendations for Cardiovascular Health and Disease Surveillance for 2030 and Beyond: A Policy Statement From the American Heart Association. Circulation. 2020;141:e104–e119.

2. Joo H, Fang J, Losby JL and Wang G. Cost of informal caregiving for patients with heart failure. Am Heart J. 2015;169:142–48 e2.

3. Bhatt AS, Abraham WT, Lindenfeld J, Bristow M, Carson PE, Felker GM, Fonarow GC, Greene SJ, Psotka MA, Solomon SD, Stockbridge N, Teerlink JR, Vaduganathan M, Wittes J, Fiuzat M, O’Connor CM and Butler J. Treatment of HF in an Era of Multiple Therapies: Statement From the HF Collaboratory. JACC Heart Fail. 2021;9:1–12.

4. Allen LA, Stevenson LW, Grady KL, Goldstein NE, Matlock DD, Arnold RM, Cook NR, Felker GM, Francis GS, Hauptman PJ, Havranek EP, Krumholz HM, Mancini D, Riegel B, Spertus JA, American Heart A, Council on Quality of C, Outcomes R, Council on Cardiovascular N, Council on Clinical C, Council on Cardiovascular R, Intervention, Council on Cardiovascular S and Anesthesia. Decision making in advanced heart failure: a scientific statement from the American Heart Association. Circulation. 2012;125:1928–52.

5. Heidenreich PA, Bozkurt B, Aguilar D, Allen LA, Byun JJ, Colvin MM, Deswal A, Drazner MH, Dunlay SM, Evers LR, Fang JC, Fedson SE, Fonarow GC, Hayek SS, Hernandez AF, Khazanie P, Kittleson MM, Lee CS, Link MS, Milano CA, Nnacheta LC, Sandhu AT, Stevenson LW, Vardeny O, Vest AR and Yancy CW. 2022 AHA/ACC/HFSA Guideline for the Management of Heart Failure: Executive Summary: A Report of the American College of Cardiology/American Heart Association Joint Committee on Clinical Practice Guidelines. J Am Coll Cardiol. 2022;79:1757–1780.

6. Levine DA, Galecki AT, Langa KM, Unverzagt FW, Kabeto MU, Giordani B and Wadley VG. Trajectory of Cognitive Decline After Incident Stroke. JAMA. 2015;314:41–51.

7. Levine DA, Gross AL, Briceno EM, Tilton N, Giordani BJ, Sussman JB, Hayward RA, Burke JF, Hingtgen S, Elkind MSV, Manly JJ, Gottesman RF, Gaskin DJ, Sidney S, Sacco RL, Tom SE, Wright CB, Yaffe K and Galecki AT. Sex Differences in Cognitive Decline Among US Adults. JAMA Netw Open. 2021;4:e210169.

8. Levine DA, Gross AL, Briceno EM, Tilton N, Kabeto MU, Hingtgen SM, Giordani BJ, Sussman JB, Hayward RA, Burke JF, Elkind MSV, Manly JJ, Moran AE, Kulick ER, Gottesman RF, Walker KA, Yano Y, Gaskin DJ, Sidney S, Yaffe K, Sacco RL, Wright CB, Roger VL, Allen NB and Galecki AT. Association Between Blood Pressure and Later-Life Cognition Among Black and White Individuals. JAMA Neurol. 2020;77:810–819.

9. Johansen MC, Ye W, Gross A, Gottesman RF, Han D, Whitney R, Briceno EM, Giordani BJ, Shore S, Elkind MSV, Manly JJ, Sacco RL, Fohner A, Griswold M, Psaty BM, Sidney S, Sussman J, Yaffe K, Moran AE, Heckbert S, Hughes TM, Galecki A and Levine DA. Association Between Acute Myocardial Infarction and Cognition. JAMA Neurol. 2023;80:723–731.

10. Gure TR, Blaum CS, Giordani B, Koelling TM, Galecki A, Pressler SJ, Hummel SL and Langa KM. Prevalence of cognitive impairment in older adults with heart failure. J Am Geriatr Soc. 2012;60:1724–9.

11. Dodson JA, Truong TT, Towle VR, Kerins G and Chaudhry SI. Cognitive impairment in older adults with heart failure: prevalence, documentation, and impact on outcomes. Am J Med. 2013;126:120–6.

12. Sterling MR, Jannat-Khah D, Bryan J, Banerjee S, McClure LA, Wadley VG, Unverzagt FW, Levitan EB, Goyal P, Peterson JC, Manly JJ, Levine DA and Safford MM. The Prevalence of Cognitive Impairment Among Adults With Incident Heart Failure: The "Reasons for Geographic and Racial Differences in Stroke" (REGARDS) Study. J Card Fail. 2019;25:130–136.

13. Qiu C, Winblad B, Marengoni A, Klarin I, Fastbom J and Fratiglioni L. Heart failure and risk of dementia and Alzheimer disease: a population-based cohort study. Arch Intern Med. 2006;166:1003–8.

14. Tilvis RS, Kahonen-Vare MH, Jolkkonen J, Valvanne J, Pitkala KH and Strandberg TE. Predictors of cognitive decline and mortality of aged people over a 10-year period. J Gerontol A Biol Sci Med Sci. 2004;59:268–74.

15. Hammond CA, Blades NJ, Chaudhry SI, Dodson JA, Longstreth WT, Jr., Heckbert SR, Psaty BM, Arnold AM, Dublin S, Sitlani CM, Gardin JM, Thielke SM, Nanna MG, Gottesman RF, Newman AB and Thacker EL. Long-Term Cognitive Decline After Newly Diagnosed Heart Failure: Longitudinal Analysis in the CHS (Cardiovascular Health Study). Circ Heart Fail. 2018;11:e004476.

16. Mohebi R, Chen C, Ibrahim NE, McCarthy CP, Gaggin HK, Singer DE, Hyle EP, Wasfy JH and Januzzi JL, Jr. Cardiovascular Disease Projections in the United States Based on the 2020 Census Estimates. J Am Coll Cardiol. 2022;80:565–578.

17. Virani SS, Alonso A, Benjamin EJ, Bittencourt MS, Callaway CW, Carson AP, Chamberlain AM, Chang AR, Cheng S, Delling FN, Djousse L, Elkind MSV, Ferguson JF, Fornage M, Khan SS, Kissela BM, Knutson KL, Kwan TW, Lackland DT, Lewis TT, Lichtman JH, Longenecker CT, Loop MS, Lutsey PL, Martin SS, Matsushita K, Moran AE, Mussolino ME, Perak AM, Rosamond WD, Roth GA, Sampson UKA, Satou GM, Schroeder EB, Shah SH, Shay CM, Spartano NL, Stokes A, Tirschwell DL, VanWagner LB, Tsao CW, American Heart Association Council on E, Prevention Statistics C and Stroke Statistics S. Heart Disease and Stroke Statistics-2020 Update: A Report From the American Heart Association. Circulation. 2020;141:e139–e596.

18. Wright JD, Folsom AR, Coresh J, Sharrett AR, Couper D, Wagenknecht LE, Mosley TH, Jr., Ballantyne CM, Boerwinkle EA, Rosamond WD and Heiss G. The ARIC (Atherosclerosis Risk In Communities) Study: JACC Focus Seminar 3/8. J Am Coll Cardiol. 2021;77:2939–2959.

19. Friedman GD, Cutter GR, Donahue RP, Hughes GH, Hulley SB, Jacobs DR, Jr., Liu K and Savage PJ. CARDIA: study design, recruitment, and some characteristics of the examined subjects. J Clin Epidemiol. 1988;41:1105–16.

20. Fried LP, Borhani NO, Enright P, Furberg CD, Gardin JM, Kronmal RA, Kuller LH, Manolio TA, Mittelmark MB, Newman A and, et al. The Cardiovascular Health Study: design and rationale. Ann Epidemiol. 1991;1:263–76.

21. Feinleib M, Kannel WB, Garrison RJ, McNamara PM and Castelli WP. The Framingham Offspring Study. Design and preliminary data. Prev Med. 1975;4:518–25.

22. Sacco RL, Boden-Albala B, Gan R, Chen X, Kargman DE, Shea S, Paik MC and Hauser WA. Stroke incidence among white, black, and Hispanic residents of an urban community: the Northern Manhattan Stroke Study. Am J Epidemiol. 1998;147:259–68.

23. The Framingham Study Criteria For Events (Sequence of Events File "SOE"). https://biolincc.nhlbi.nih.gov/media/studies/framoffspring/Protocols/Criteria%20for%20Events%20-%20Sequence%20of%20Events%20File%20SOE.pdf?link_time=2022-11-14_23:11:26.747440.

24. Bahrami H, Bluemke DA, Kronmal R, Bertoni AG, Lloyd-Jones DM, Shahar E, Szklo M and Lima JA. Novel metabolic risk factors for incident heart failure and their relationship with obesity: the MESA (Multi-Ethnic Study of Atherosclerosis) study. J Am Coll Cardiol. 2008;51:1775–83.

25. Bibbins-Domingo K, Pletcher MJ, Lin F, Vittinghoff E, Gardin JM, Arynchyn A, Lewis CE, Williams OD and Hulley SB. Racial differences in incident heart failure among young adults. N Engl J Med. 2009;360:1179–90.

26. Rosamond WD, Chang PP, Baggett C, Johnson A, Bertoni AG, Shahar E, Deswal A, Heiss G and Chambless LE. Classification of heart failure in the atherosclerosis risk in communities (ARIC) study: a comparison of diagnostic criteria. Circ Heart Fail. 2012;5:152–9.

27. CHS: Event Data Collection Overview. https://chs-nhlbi.org/EventsData.

28. Rincon F, Dhamoon M, Moon Y, Paik MC, Boden-Albala B, Homma S, Di Tullio MR, Sacco RL and Elkind MS. Stroke location and association with fatal cardiac outcomes: Northern Manhattan Study (NOMAS). Stroke. 2008;39:2425–31.

29. Camplain R, Kucharska-Newton A, Loehr L, Keyserling TC, Layton JB, Wruck L, Folsom AR, Bertoni AG and Heiss G. Accuracy of Self-Reported Heart Failure. The Atherosclerosis Risk in Communities (ARIC) Study. J Card Fail. 2017;23:802–808.

30. Hachinski V, Iadecola C, Petersen RC, Breteler MM, Nyenhuis DL, Black SE, Powers WJ, DeCarli C, Merino JG, Kalaria RN, Vinters HV, Holtzman DM, Rosenberg GA, Wallin A, Dichgans M, Marler JR and Leblanc GG. National Institute of Neurological Disorders and Stroke-Canadian Stroke Network vascular cognitive impairment harmonization standards. Stroke. 2006;37:2220–41.

31. Manly JJ, Schupf N, Stern Y, Brickman AM, Tang MX and Mayeux R. Telephone-based identification of mild cognitive impairment and dementia in a multicultural cohort. Arch Neurol. 2011;68:607–14.

32. Briceno EM, Gross AL, Giordani BJ, Manly JJ, Gottesman RF, Elkind MSV, Sidney S, Hingtgen S, Sacco RL, Wright CB, Fitzpatrick A, Fohner AE, Mosley TH, Yaffe K and Levine DA. Pre-Statistical Considerations for Harmonization of Cognitive Instruments: Harmonization of ARIC, CARDIA, CHS, FHS, MESA, and NOMAS. J Alzheimers Dis. 2021;83:1803–1813.

33. Muthen LK and Muthen BO. Mplus user’s guide: Eighth Edition. Los Angeles, CA.

34. Asparouhov T and Muthén B. Plausible values for latent variables using Mplus. Technical Report. http://www.statmodel.com/download/Plausible.pdf. 2010;2022.

35. Chen LY, Norby FL, Gottesman RF, Mosley TH, Soliman EZ, Agarwal SK, Loehr LR, Folsom AR, Coresh J and Alonso A. Association of Atrial Fibrillation With Cognitive Decline and Dementia Over 20 Years: The ARIC-NCS (Atherosclerosis Risk in Communities Neurocognitive Study). J Am Heart Assoc. 2018;7.

36. Mann DL. Innate immunity and the failing heart: the cytokine hypothesis revisited. Circ Res. 2015;116:1254–68.

37. Iadecola C and Gorelick PB. Converging pathogenic mechanisms in vascular and neurodegenerative dementia. Stroke. 2003;34:335–7.

38. Doyle KP, Quach LN, Sole M, Axtell RC, Nguyen TV, Soler-Llavina GJ, Jurado S, Han J, Steinman L, Longo FM, Schneider JA, Malenka RC and Buckwalter MS. B-lymphocyte-mediated delayed cognitive impairment following stroke. J Neurosci. 2015;35:2133–45.

39. Villringer A and Laufs U. Heart failure, cognition, and brain damage. Eur Heart J. 2021;42:1579–1581.

40. Glymour MM and Manly JJ. Lifecourse social conditions and racial and ethnic patterns of cognitive aging. Neuropsychol Rev. 2008;18:223–54.

41. Lam CSP, Arnott C, Beale AL, Chandramouli C, Hilfiker-Kleiner D, Kaye DM, Ky B, Santema BT, Sliwa K and Voors AA. Sex differences in heart failure. Eur Heart J. 2019;40:3859–3868c.

42. Mielke MM, Vemuri P and Rocca WA. Clinical epidemiology of Alzheimer’s disease: assessing sex and gender differences. Clin Epidemiol. 2014;6:37–48.

43. Wolinsky FD, Unverzagt FW, Smith DM, Jones R, Stoddard A and Tennstedt SL. The ACTIVE cognitive training trial and health-related quality of life: protection that lasts for 5 years. J Gerontol A Biol Sci Med Sci. 2006;61:1324–9.

44. Riegel B, Moser DK, Anker SD, Appel LJ, Dunbar SB, Grady KL, Gurvitz MZ, Havranek EP, Lee CS, Lindenfeld J, Peterson PN, Pressler SJ, Schocken DD, Whellan DJ, American Heart Association Council on Cardiovascular N, American Heart Association Council on Cardiovascular N, American Heart Association Council on Clinical C, American Heart Association Council on Nutrition PA, Metabolism, American Heart Association Interdisciplinary Council on Quality of C and Outcomes R. State of the science: promoting self-care in persons with heart failure: a scientific statement from the American Heart Association. Circulation. 2009;120:1141–63.

45. Unutzer J, Katon W, Callahan CM, Williams JW, Jr., Hunkeler E, Harpole L, Hoffing M, Della Penna RD, Noel PH, Lin EH, Tang L and Oishi S. Depression treatment in a sample of 1,801 depressed older adults in primary care. J Am Geriatr Soc. 2003;51:505–14.

46. Clark LJ, Gatz M, Zheng L, Chen YL, McCleary C and Mack WJ. Longitudinal verbal fluency in normal aging, preclinical, and prevalent Alzheimer’s disease. Am J Alzheimers Dis Other Demen. 2009;24:461–8.

47. Bennett HP, Corbett AJ, Gaden S, Grayson DA, Kril JJ and Broe GA. Subcortical vascular disease and functional decline: a 6-year predictor study. J Am Geriatr Soc. 2002;50:1969–77.

48. Petersen RC, Lopez O, Armstrong MJ, Getchius TSD, Ganguli M, Gloss D, Gronseth GS, Marson D, Pringsheim T, Day GS, Sager M, Stevens J and Rae-Grant A. Practice guideline update summary: Mild cognitive impairment: Report of the Guideline Development, Dissemination, and Implementation Subcommittee of the American Academy of Neurology. Neurology. 2018;90:126–135.

49. Verhaeghen P, Marcoen A and Goossens L. Improving memory performance in the aged through mnemonic training: a meta-analytic study. Psychol Aging. 1992;7:242–51.

50. Sbolli M, Fiuzat M, Cani D and O’Connor CM. Depression and heart failure: the lonely comorbidity. Eur J Heart Fail. 2020;22:2007–2017.

